# Prevalence, Correlates, and Time-to-Event Analysis of Female Sterilisation Failure in India: A Nationally Representative Study Using NFHS-5 Data

**DOI:** 10.64898/2026.08.03.26359553

**Authors:** Pavan Pandey

## Abstract

**Background:** Female sterilisation is the most widely used contraceptive method in India. But nationally representative evidence on the frequency, outcome and correlates of sterilisation failure remains scarce. The objectives of this study were to estimate the prevalence of contraceptive failure, describe the outcomes of pregnancies resulting from failure, determine the time interval between sterilisation and failure, and compute the Pearl Index.

**Methods:** This secondary data analysis used the Individual Recode dataset of the National Family Health Survey-5 (NFHS-5, 2019–21). Sterilisation failure was derived from reproductive and contraceptive history variables. Weighted prevalence, socio-demographic and procedural correlates, and a Pearl Index were estimated using survey-adjusted statistical methods.

**Results:** Among 189,021 women who had adopted sterilisation, 623 (weighted prevalence 0.34%) experienced a failure. The most common outcome of sterilisation failure was abortion (96.0%). The Pearl Index was 2.87 failures per 100 woman-years (95% CI: 2.58–3.20). Failure was significantly associated with years since sterilisation, decade and age at sterilisation, marital duration, parity, place and type of sterilisation (all p<0.05), but not with education, religion, caste, or wealth index. Failures occurred at any point from one month to over a decade after the procedure, with a disproportionately higher share among women sterilised at mobile clinics/camps.

**Conclusion:** Female sterilisation failure in India, while uncommon, is not negligible and persists well beyond the immediate postoperative period. Procedural quality of care, particularly at camp-based settings, appears to be a key modifiable driver. The postoperative counselling needs to include a clear message that a missed period any time after (months to year) sterilisation can be a pregnancy resulting from failure of sterilization.

**Key Messages:** 

**What is already known on this topic:** – Female sterilisation is the most widely used contraceptive method in India. Existing evidence on sterilisation failure is drawn almost entirely from small, single-centre hospital case series that lack a defined denominator.

**What this study adds:** – Using nationally representative NFHS-5 data, this study provides the first national estimate of female sterilisation failure (prevalence 0.34%; Pearl Index 2.87 per 100 woman-years). Sterilisation failure can occur at any point from one month to over two decades after the procedure and is disproportionately concentrated among women sterilised at mobile clinics/camps.

**How this study might affect research, practice, or policy:** – These findings support strengthening quality assurance at camp-based sterilisation services. Additionally, we need to revise post-sterilisation counselling and certification to clearly communicate that a missed period any time after sterilization can be an unintended pregnancy as a result of failure.

## Introduction

Globally, female sterilisation is one of the most widely used methods of contraception.^1^ Sterilisation is a simple, one-time, and cost-effective procedure that requires minimal follow-up care. Once completed, it does not depend on user’s or partner’s compliance.^1^ These features make sterilisation more user-friendly than other modern contraceptive methods. With the refinement of surgical techniques and safer anaesthetic practices, postoperative morbidity and mortality associated with sterilisation have been reduced to minimum.^2,3^ It is probably secondary to these advantages; female sterilisation has remained the most preferred method of contraception in India for several decades.^4^ According to the National Family Health Survey (NFHS-4, NFHS-5 and NFHS-6), sterilisation was used by 36%, 38% and 36.5% of married women, respectively.^5,6^ In absolute terms, about four million sterilisation procedures are performed annually in India.^7^

Given its popularity and widespread acceptance among Indian couples, and its permanent nature, it is essential to have a comprehensive understanding of the short- and long-term reproductive and demographic implications of this procedure. One important aspect among all post-sterilisation events is the failure of sterilisation. Although sterilisation is promoted as a one-time permanent procedure, pregnancy after sterilisation, though uncommon, is still possible.^8^ A failed sterilisation can result in an unintended pregnancy, which may lead to a live birth, miscarriage, or voluntary abortion. Each of these events can be a source of emotional turmoil and economic stress for affected couples. In some cases, sterilisation failure can also invite legal action against the surgeon or hospital.^9^ In India, sterilisation failure also has financial implications for the Government, as couples who experience sterilisation failure are eligible for financial compensation under the Family Planning Indemnity Scheme (FPIS).^10^ Reliable estimates of sterilisation failure are, therefore, essential for strengthening pre- and post-operative counselling, and protecting the rights of both clients and providers.

Despite the vast scale of India’s sterilisation programme, empirical evidence on the prevalence and fate of sterilisation failure remains very limited. Only a few studies have examined the prevalence of sterilisation failure, and most such studies are based on very small-scale, single-centre data. To the best of the authors’ knowledge, no recent, national-level study has estimated rates of female sterilisation failure in India. We, therefore, undertook this secondary data analysis of NFHS-5 data with the following objectives:

1. To estimate the prevalence of contraceptive failure after adopting female sterilisation.
2. To determine the outcomes of pregnancies resulting from failed sterilisation: live birth, abortion, or current pregnancy.
3. To determine the time interval between sterilisation and the subsequent failure event.
4. To compute the Pearl Index for female sterilisation based on NFHS-5 data.

## Methodology

### Survey Details

This study is a secondary data analysis based on the nationally representative data collected for the National Family Health Survey–5 (NFHS-5). The NFHS-5 fieldwork was conducted in two phases: Phase I (June 2019–January 2020) covered 17 states and 5 union territories, and Phase II (January 2020–April 2021) covered 11 states and 3 union territories. The NFHS series, initiated in 1992–93, is designed to collect reliable and national-level data on population, health, and nutrition. The NFHS is the Indian version of the global Demographic and Health Surveys (DHS). The NFHS-5 survey was implemented by the International Institute for Population Sciences (IIPS), Mumbai with technical assistance from ICF International, USA, through the Demographic and Health Surveys (DHS) Programme, funded by USAID.

### Survey Design

A stratified two-stage sampling design was adopted for NFHS-5. In the first stage, the Primary Sampling Units (PSUs) were selected: villages in rural areas and Census Enumeration Blocks (CEBs) in urban areas. In the second stage, in every selected rural and urban cluster, 22 households were randomly selected with systematic sampling. Data collection was carried out by 17 field agencies, resulting in successful interviews from 636,699 households, 724,115 women aged 15–49 years, and 101,839 men aged 15–54 years.

### Study Population

The study population included all surveyed women who reported using female sterilisation as their current method of contraception (n = 189,021). Women who reported using other methods of contraception including male sterilisation were excluded. The analysis was based on the Individual Recode (IR) dataset of NFHS-5, which contains detailed information on women’s demographic, reproductive, and contraceptive characteristics.

### Definition of Sterilisation Failure

The NFHS-5 dataset does not have a readymade variable indicating sterilisation failure. Therefore, the failure was derived using logical conditions based on available reproductive and contraceptive history of women. A woman was classified as having experienced a sterilisation failure if any one of the following conditions was met:

(i) The date of birth of the last or youngest child was later than the reported date of sterilisation, OR
(ii) The date of the most recent abortion was later than the date of sterilisation, OR
(iii) She reported adopting sterilisation in the past but was pregnant at the time of the survey.

These criteria were developed using the following variables available in the NFHS-5 datasets: pregnant at the time of interview (v325); current method of contraception (v312); month and year of sterilisation (v315 and v316); month and year of birth of youngest child (b1_01, b2_01), and month and year of most recent abortion (v229, v230). Thereafter, a composite binary outcome variable, “failure of sterilisation,” was generated where 1 indicated any form of failure and 0 indicated no failure.

### Independent Variables

The main independent variables were: Age at sterilisation (v320); Years since sterilisation (v319); Parity at sterilisation (v322); Type of healthcare facility for sterilisation (v326); Residence (urban or rural) (v025); socio-demographic variables (woman’s current age, education (v106), caste (v131), religion (v130), and wealth index (v190)).

### Statistical Analysis

All analyses were conducted using Stata version 17.0 (personal license). The complex sampling design of NFHS-5 was accounted for by applying sampling weights (v005/1,000,000) and the survey design variables (primary sampling unit: v021; strata: v023). Frequencies and percentages were computed for categorical variables and means, and standard deviations were calculated for continuous variables. The weighted prevalence of sterilisation failure was estimated among all sterilized women using the “svy” and “subpop” commands to produce nationally representative estimates. The association between sterilisation failure and selected independent variables was examined using cross-tabulations and Pearson’s chi-square / Fisher’s exact tests. The mean time from sterilisation to the occurrence of a failure event was calculated. The Pearl Index was also computed for female sterilisation to measure the number of failures per 100 sterilisation procedures per year of exposure.

### Ethical Considerations

Ethical clearance was not required for this study as it was a secondary data analysis of a de-identified, open-access dataset. NFHS-5 data was downloaded from DHS programme’s website after approval (https://dhsprogram.com/methodology/survey/survey-display-541.cfm). The original survey protocol received ethical clearance from the Institutional Review Board (IRB) of IIPS, Mumbai, and the ICF Institutional Review Board, USA. The details of the survey can be found elsewhere.^6^

## Results

**Fig 1:**
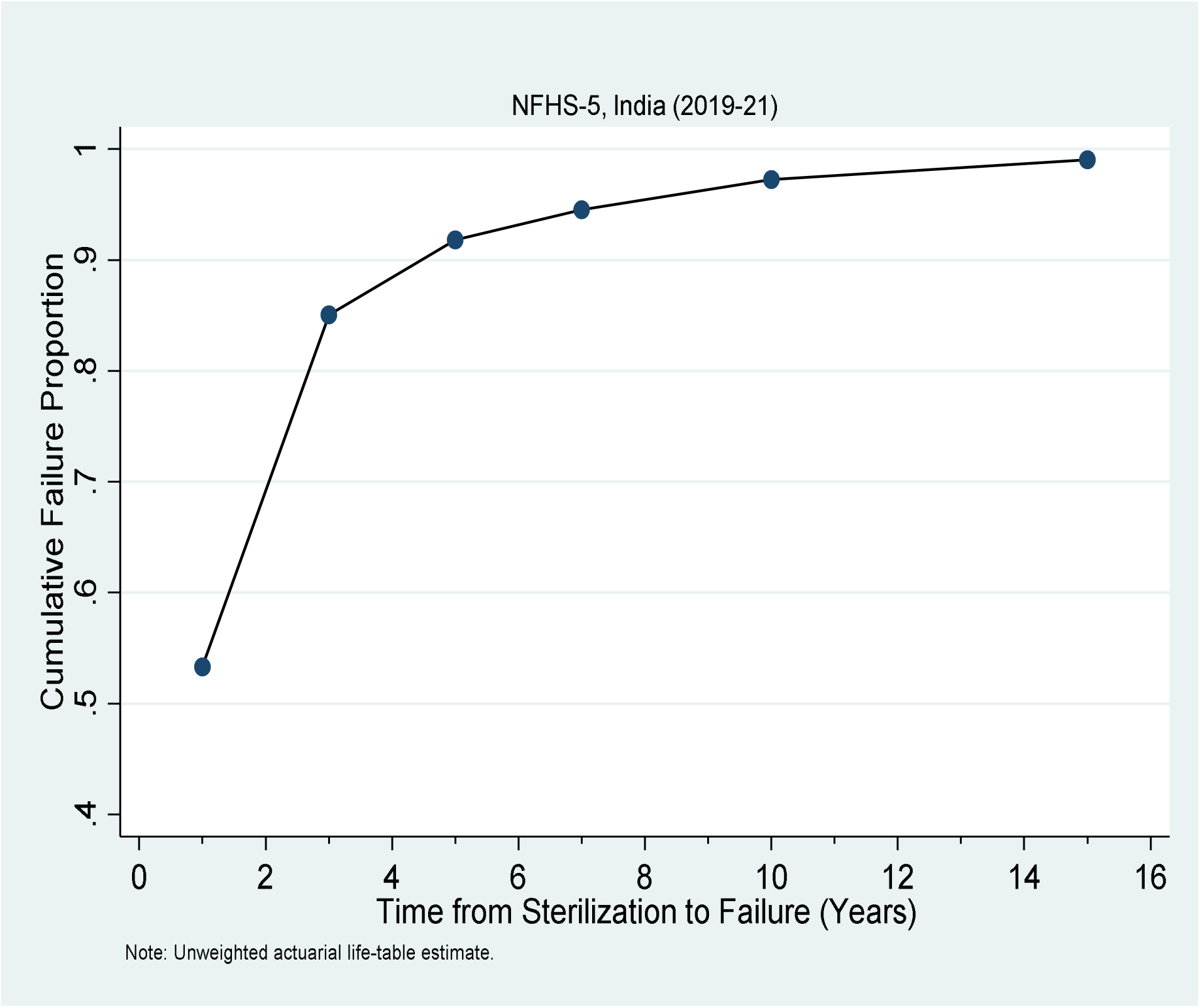
Cumulative Failure of Female Sterilization

Table 1 illustrates that among 189,021 women who reported adopting sterilisation in NFHS-5, only 623 women experienced sterilisation failure (weighted prevalence 0.34%). Among these 623 women, the pregnancy resulting from failed female sterilisation was aborted in 589 cases (96.0%) while 34 women were still pregnant at the time of the survey (4%), and there were no cases of a live birth resulting from a failed sterilisation procedure. The time difference between sterilization procedure and failure event ranged from a minimum of one month to twenty-one years. The Pearl Index was 2.87 failures per 100 woman-years (95% CI: 2.58–3.20).

**Table 1.** Prevalence of sterilization failure and outcome type among sterilized women (N = 189,021)

| Characteristic | n (weighted %) |
| --- | --- |
| ***Sterilization failure status*** |  |
| No failure | 188,398 (99.66) |
| Failed | 623 (0.34) |
| ***Outcome of failed sterilization (% of all failed sterilized cases)*** |  |
| Abortion | 589 (96.0) |
| Untermiated pregnancy | 34 (4.0) |
| Live Birth | 0 (0.0) |
| ***Time to failure event (among n=623 failures)*** |  |
| < 6 months | 205 (32.26) |
| 6-12 months | 127 (22.28) |
| 1-2 years | 143 (23.05) |
| > 2 years | 148 (22.40) |
| Range | 1 month – 21 years |
| **Pearl Index (per 100 woman-years)** | 2.87 (95% CI: 2.58-3.20) |

*Note: n = unweighted frequency; % = weighted percentage.*

Table 2 shows the socio-demographic parameters of sterilized women stratified by failure. Women with sterilisation failure were older than those without failure, with a mean age of 40.58 years compared with 37.73 years. Failure was significantly associated with age group (p<0.001) and place of residence (p=0.042). Educational level, religion, caste or tribe category, and wealth index were not significantly associated with sterilisation failure.

**Table 2.** Socio-demographic characteristics of sterilized women, by sterilization failure status.

| Characteristic | No failure,<br>n (weighted %) | Failed,<br>n (weighted %) | P-value |
| --- | --- | --- | --- |
| ***Age at time of Survey (years)*** |  |  | **<0.001** |
| 15-24 | 4,613 (2.74) | 9 (0.82) |  |
| 25-34 | 56,197 (30.48) | 110 (15.09) |  |
| 35-49 | 127,588 (66.79) | 504 (84.09) |  |
| Mean, SD | 37.73 (37.68-37.77) | 40.58 (39.97-41.19) |  |
| ***Type of place of residence*** |  |  | **0.042** |
| Urban | 42,655 (30.33) | 160 (35.95) |  |
| Rural | 145,743 (69.67) | 463 (64.05) |  |
| ***Highest educational level*** |  |  | **0.378** |
| No education | 73,043 (35.76) | 225 (34.29) |  |
| Primary | 32,082 (17.02) | 131 (19.96) |  |
| Secondary | 73,627 (40.89) | 245 (40.97) |  |
| Higher | 9,646 (6.34) | 22 (4.78) |  |
| ***Religion*** |  |  | **0.177** |
| Hindu | 162,469 (87.69) | 538 (90.77) |  |
| Muslim | 12,557 (7.52) | 42 (5.68) |  |
| Other | 13,372 (4.79) | 43 (3.55) |  |
| **Caste/tribe category** |  |  | **0.964** |
| Caste | 162,873 (90.11) | 540 (90.51) |  |
| Tribe | 19,419 (6.22) | 61 (5.91) |  |
| No caste/tribe | 6,106 (3.67) | 22 (3.58) |  |
| **Wealth index** |  |  | **0.611** |
| Poorest | 35,959 (17.03) | 107 (14.14) |  |
| Poorer | 42,027 (20.37) | 135 (21.19) |  |
| Middle | 44,180 (23.12) | 150 (22.34) |  |
| Richer | 38,892 (22.19) | 132 (24.11) |  |
| Richest | 27,340 (17.28) | 99 (18.22) |  |
| *Note: n = unweighted frequency; % = weighted column percentage* |  |  |  |
| *P-values are from the design-based F-test (Rao-Scott corrected Pearson chi-square).* |  |  |  |

Table 3 shows the sterilisation-related parameters among the surveyed women stratified by failure. The data analysis showed that sterilisation failure was significantly associated with years since sterilisation, decade of sterilisation, age at sterilisation, marital duration at sterilisation, parity, place of sterilisation, and regret following sterilisation.

**Table 3.** Sterilization-related characteristics of sterilized women, by sterilization failure status.

| Characteristic | No failure, n<br>(weighted %) | Failed, n<br>(weighted %) | P-value |
| --- | --- | --- | --- |
| ***Years since sterilization*** |  |  | **<0.001** |
| < 4 years | 29,280 (15.73) | 25 (3.11) |  |
| 4-7 years | 35,769 (18.53) | 60 (8.20) |  |
| ≥ 8 years | 123,349 (65.74) | 538 (88.69) |  |
| Mean, SD | 11.75 (11.70-11.80) | 14.85 (14.20-15.50) |  |
| ***Decade of sterilization*** |  |  | **<0.001** |
| Before 1990s | 134 (0.10) | 0 (0.00) |  |
| 1990s | 23,189 (13.77) | 101 (20.34) |  |
| 2000s | 78,543 (41.43) | 350 (56.09) |  |
| 2010 onward | 86,532 (44.69) | 172 (23.56) |  |
| ***Age at sterilization*** |  |  | **0.022** |
| < 25 years | 75,594 (43.11) | 265 (42.18) |  |
| 25-29 years | 70,179 (36.13) | 242 (42.15) |  |
| 30-34 years | 31,466 (15.57) | 88 (12.70) |  |
| ≥ 35 years | 11,159 (5.19) | 28 (2.97) |  |
| Mean, SD | 25.98 (25.94-26.01) | 25.73 (25.35-26.10) |  |
| ***Marital duration at sterilization*** |  |  | **0.001** |
| Before marriage | 246 (0.15) | 8 (0.70) |  |
| < 10 years | 130,577 (71.41) | 444 (73.57) |  |
| 10-19 years | 53,736 (26.58) | 164 (25.14) |  |
| ≥ 20 years | 3,768 (1.86) | 7 (0.60) |  |
| **Parity at sterilization** |  |  | **<0.001** |
| 0 | 174 (0.10) | 8 (0.70) |  |
| 1 | 5,671 (3.43) | 58 (10.53) |  |
| 2 | 80,025 (45.98) | 226 (39.72) |  |
| 3 | 57,383 (28.97) | 192 (28.38) |  |
| 4+ | 45,145 (21.52) | 139 (20.67) |  |
| **Place of sterilization** |  |  | **<0.001** |
| District / higher govt.<br>hospital | 85,229 (43.53) | 284 (43.13) |  |
| CHC / PHC / SC | 66,274 (34.33) | 177 (27.19) |  |
| Mobile clinic / camp | 7,738 (3.24) | 59 (8.89) |  |
| Private / NGO / other | 29,157 (18.89) | 103 (20.80) |  |
| **Regrets sterilization** |  |  | **0.016** |
| No | 178,101 (95.08) | 548 (92.14) |  |
| Yes | 9,877 (4.92) | 43 (7.86) |  |
| **Type of Sterilization** |  |  | **0.003** |
| Interval | 136474 (68.0) | 485 (75.9) |  |
| Postpartum | 51750 (32.0) | 130 (24.1) |  |

| Characteristic | No failure, n<br>(weighted %) | Failed, n<br>(weighted %) | P-value |
| --- | --- | --- | --- |

*Note: n = unweighted frequency; % = weighted column percentage*
*P-values are from the design-based F-test (Rao-Scott corrected Pearson chi-square).*

## Discussion

This secondary data analysis of NFHS-5 found that among 189,021 women who had adopted female sterilisation, 0.34% (unweighted n =623) experienced a subsequent contraceptive failure. The most common outcome of a failed sterilisation was an induced abortion (96.0%). The Pearl Index for female sterilisation was 2.87 failures per 100 woman-years (95% CI: 2.58– 3.20). These findings have implications for clinical counselling, service delivery, and family planning programme, both in India and elsewhere.

Most empirical evidence from India on sterilisation failure is derived from small-scale, hospital-based studies (mostly descriptive case series) conducted at a single institution. In all such studies, women were identified after they presented with sterilisation failure and there was no defined denominator of all the women sterilised at that facility over a given time period.^8,11–13^ Such studies, at best, can only describe the clinical and demographic profile of women who experienced failure. Such studies cannot estimate the prevalence of sterilisation failure. In all these aspects, the present study is different from other studies conducted earlier. In addition, this study draws on a nationally representative, population-based sample. This allowed sterilization failure to be expressed as a true prevalence, and as a formal Pearl Index. Hospital-based studies simply cannot produce these figures, regardless of their size or how long it followed the sterilised women. To the authors’ knowledge, this is the first study to collectively integrate five distinct elements within a single analysis: a nationally representative sample (in contrast to a single institute), overall prevalence of sterilization failure, the specific reproductive outcome of each failure, the time elapsed between sterilisation and failure, and a Pearl Index. Additionally, this study also provides a wider set of socio-demographic and procedural correlates of sterilisation failure. This allows individual-level factors, such as age and parity, to be examined alongside health-system-level factors, such as place of sterilisation, within the same analysis. This combination was not previously available in the Indian literature on this subject.

The prevalence and rate estimates from this study can be compared against three bodies of existing evidence. Internationally, the US Collaborative Review of Sterilization (CREST) followed a cohort of 10,685 women for up to 14 years. It reported a 10-year cumulative failure rate of 1.85%.^14,15^ The failure rate in that study varied substantially by technique.^14^ Trussell et al., have reported a conservative, typical-use failure probability of female sterilisation at approximately 0.5%.^16^ The government of India’s (GoI) official guidelines on female sterilization suggest a failure rate of less than 1 pregnancy per 100 women (5 per 1,000) in the first year after sterilisation, rising to approximately 2 pregnancies per 100 women (18–19 per 1,000) over 10 years.^17^ These figures imply an average annual rate of around 0.2 per 100 woman-years. This is appreciably lower than the 2.87 per 100 woman-years observed in the present study. Smaller Indian hospital-based case series are difficult to compare directly, since they lack a clear denominator. Even so, they have suggested failure rates in the range of 0.1–0.8%. ^8^ Our national prevalence estimate of 0.34% is broadly consistent with the range reported by small Indian case series. However, our Pearl Index is appreciably higher than both CREST-era Western estimates and India’s own programmatic figures.

Among the sterilisation related parameter, place of sterilisation was significantly associated with failure (p <0.001). Mobile camp sterilisation accounted for a markedly higher share of failures (8.89%) than their share of the no-failure group (3.24%). This finding is indirectly supported by several other independent studies which have reported that compliance to every aspect of sterilisation procedure viz. preoperative, intraoperative, and postoperative protocols is compromised in the camp setting.^18–22^ Every year, a significant share of India’s 4 million sterilisation are conducted at such high-throughput and time-constrained sterilisation camps. This is not the case in western countries; hence, the sub-standard surgical practices followed at sterilisation camps may plausibly explain why India’s national Pearl Index in this study exceeds Western cohort-based estimates.

Among the various factors associated with sterilisation failure, the most consistent pattern was the relationship with the time elapsed since the procedure. This is reflected in each of the variables influenced by the ‘time difference’ between the date of sterilisation and date of interview e.g., current age, decade of sterilisation, years since sterilisation etc. We observed that women with failure had a substantially longer mean duration since sterilisation than those without failure — 14.85 years compared with 11.75 years. Among women with failure, 88.69% had been sterilised eight or more years earlier. Among women without failure, this figure was only 65.74%. The decade-of-sterilisation results reinforce this pattern. Women sterilised in the 1990s and 2000s accounted for a disproportionately large share of failures — 20.34% and 56.09% respectively — compared with their share of the no-failure group. This mirrors a finding from the CREST study: the risk of pregnancy after tubal sterilisation persists throughout a woman’s reproductive years. It is not confined to an initial high-risk period. ^14,15^ Furthermore, our analysis showed that 32.26% of failures occurred within six months, and a further 22.28% occurred within the first year. This suggests that a meaningful share of failures in this population resulted from incomplete occlusion at the time of the original procedure, rather than from late recanalization alone. Age at sterilisation (p=0.022) and marital duration at sterilisation (p=0.001) followed a related pattern. This is unlikely to reflect any distinct biological vulnerability in younger women. It is more simply explained by exposure time; younger women simply have more years during which a failure could occur.

The time-to-failure findings from this study carry a direct and practical message for postoperative counselling. In India, a woman is issued a sterilisation certificate only after her immediate next menstrual period or a negative pregnancy test conducted at least 4-6 weeks after surgery (in cases of postpartum amenorrhea).^17^ There is a common perception among sterilised women and frontline healthcare workers that the window of risk closes after receiving sterilisation certificate. Our findings contradict this perception. Failure in this study occurred at virtually any point along a woman’s reproductive lifespan following sterilisation, ranging from a minimum of 1 month to 21 years after the procedure. This means sterilisation, while highly effective, does not eliminate the possibility of pregnancy entirely and this residual risk does not expire after a fixed “safe” period. Therefore, women should be clearly counselled at the time of issue of certificate: a missed menstrual period, whenever it occurs, is a reason to take a pregnancy test and thereafter contact healthcare provider. This applies whether the missed period comes months, years, or over a decade after the procedure. Under no circumstances, a woman should assume that a missed period, so long after sterilisation, cannot be pregnancy related. Early recognition of a post-sterilisation pregnancy has direct clinical significance. If the pregnancy is ectopic, a delayed diagnosis increases the risk that it will not be identified until it becomes a medical emergency. This is a recognised and potentially life-threatening complication specific to sterilisation failure.^23^ In practice, this means informed consent and counselling protocols for sterilisation in India should clearly communicate the possibility of late failure. This message should be reinforced as a clear advisory to be printed/written on the sterilisation certificate issued to the women after the procedure.

### Strengths and limitations

The two biggest strengths of present study are the large, nationally representative sample and a long duration of follow up since sterilization. To the authors’ knowledge, it represents the first and most recent national-level estimate of female sterilisation failure in India. This study also has few limitations secondary to the content and quality of NFHS-5 data. First, the estimated prevalence of sterilisation failure is completely dependent on the accuracy of the NFHS data. Several studies have pointed out the inaccuracies in DHS data.^24,25^ The dates of sterilisation, birth, and abortion in NFHS-5 are recorded only to the month and year. Hence, we have to assume all these events were conducted on the 15^th^ of every month. Secondly, the NFHS-5 survey does not record the specific surgical approach used for sterilisation — laparoscopic versus open (minilaparotomy). This is a well-established determinant of failure risk. This is a factor shown internationally, including in the CREST study, to be among the strongest determinants of failure. ^14,15^ Third, the survey does not identify ectopic pregnancy as a distinct outcome category. Future studies with facility- or provider-linked data are needed to properly characterise the true burden of post-sterilisation ectopic pregnancy in India.

## Conclusion and policy implications

This study provides the recent, nationally representative estimate of female sterilisation failure in India. It identifies place of sterilisation, and time since sterilisation as its principal correlates. The sterilisation failure cases were disproportionately higher among women sterilised in camp and mobile-clinic settings. This suggests that procedural quality, rather than the characteristics of the women themselves, is the most policy-actionable target for reducing sterilisation failure in India. This supports continued investment in quality assurance for sterilisation services, particularly at camp and mobile-clinic settings. Post-procedure counselling should also be revised. It should make clear that failure can occur at any point over the following decade or more, and that any missed period, however long after the procedure, warrants a pregnancy test and prompt contact with a healthcare provider. Sterilisation failure in India carries financial and legal implications, including eligibility for compensation under the Family Planning Indemnity Scheme. Given this, more robust prospective surveillance of sterilisation outcomes is needed. Linking this surveillance to procedure-level and provider-level data, where possible, would substantially strengthen the evidence base for both clinical counselling and programme oversight going forward.

## Programme / Policy Recommendations

**1. Strengthen quality assurance specifically at camp and mobile-clinic sterilisation services.** Given that a substantial share of India’s roughly four million annual sterilisations are performed through these camps. Ensuring compliance to prescribed preoperative, intraoperative, and postoperative protocols are likely to yield the largest reduction in national failure rates of any single lever available to the programme.
**2. Revise post-sterilisation counselling and the sterilisation certificate to explicitly communicate long-term residual risk.** Failure in this study occurred across the full span of follow-up, from within the first month to over two decades after the procedure. Counselling protocols and the certificate itself should be revised to state clearly that a missed menstrual period, whenever it occurs, warrants a pregnancy test and prompt contact with a healthcare provider.

## Data Availability

All data produced are available online at https://dhsprogram.com/methodology/survey/survey-display-541.cfm

https://dhsprogram.com/data/available-datasets.cfm

## References

1. Lawrie TA, Kulier R, Nardin JM. Techniques for the interruption of tubal patency for female sterilisation. Cochrane Database Syst Rev [Internet]. 2016;2016(8). Available from: 10.1002/14651858.cd003034.pub4

2. Kulier R, Boulvain M, Walker D, Candolle G, Campana A. Minilaparotomy and endoscopic techniques for tubal sterilisation. Cochrane database Syst Rev. 2004;2004(3):CD001328.

3. Jokinen E, Heino A, Karipohja T, Gissler M, Hurskainen R. Safety and effectiveness of female tubal sterilisation by hysteroscopy, laparoscopy, or laparotomy: a register based study. BJOG. 2017 Nov;124(12):1851–7.

4. de Oliveira IT, Dias JG, Padmadas SS. Dominance of sterilization and alternative choices of contraception in India: an appraisal of the socioeconomic impact. PLoS One. 2014;9(1):e86654.

5. National Family Health Survey - 6, 2023-24: India Country Fact Sheet [Internet]. Mumbai. India; 2026. Available from: https://www.nfhsiips.in/nfhsuser/publication.php

6. (IIPS) II for PS, ICF. National Family Health Survey (NFHS-5), 2019-21: India. Mumbai: IIPS; 2021.

7. Division S. HMIS Standard Reports: Indicator Wise- Family Planning [Internet]. 2024. Available from: https://hmis.mohfw.gov.in/#!/standardReports

8. Backialakshmi P. A comprehensive review of female sterilisation failure. Int J Med Public Heal. 2025;15(1):1645–9.

9. Argent V. Failed sterilization and the law. BJOG An Int J Obstet Gynaecol [Internet]. 1988;95(2):113–5. Available from: 10.1111/j.1471-0528.1988.tb06837.x

10. Manual for Family Planning Indemnity Scheme (2nd Edition) [Internet]. New Delhi; 2016. Available from: http://nrhm.gov.in/images/pdf/programmes/family-planing/schemes/FPIS_2nd_Edition_2016.pdf

11. Sharma R, Guleria K, Suneja A, Gupta R. Female Sterilization Failure reported during 5 Years at a Tertiary Care Teaching Hospital: A Retrospective Survey. J South Asian Fed Obstet Gynaecol [Internet]. 2017;9(3):245–9. Available from: 10.5005/jp-journals-10006-1504

12. Date S, Rokade J, Mule VD, Dandapannavar S. Female sterilization failure: Review over a decade and its clinicopathological correlation. Int J Appl Basic Med Res [Internet]. 2014;4(2):81. Available from: 10.4103/2229-516x.136781

13. Rani R, Sharma R, Kohli C. Female sterilization failure, consequences and further contraception: a review of cases over ten years. Int J Reprod Contracept Obstet Gynecol [Internet]. 2020;9(10):4032. Available from: 10.18203/2320-1770.ijrcog20204282

14. Peterson HB, Xia Z, Hughes JM, Wilcox LS, Tylor LR, Trussell J, et al. The risk of pregnancy after tubal sterilization: Findings from the U.S. collaborative review of sterilization. Am J Obstet Gynecol. 1996;174(4):1161–70.

15. Baill IC, Cullins VE, Pati S. Counseling issues in tubal sterilization. PubMed [Internet]. 2003;67(6):1287–94. Available from: https://pubmed.ncbi.nlm.nih.gov/12674457

16. Trussell J. Contraceptive failure in the United States. Contraception. 2011 May;83(5):397–404.

17. Government of India: Ministry of Health and Family Welfare. Reference Manual for Female Sterilization [Internet]. Vol. 1, Family Planning Guidelines. New Delhi; 2014. Available from: http://nhm.gov.in/images/pdf/programmes/family-planing/guidelines/Ref_Manual_for_Female_Sterilization.pdf

18. Joseph VJK, Mozumdar A, Lhungdim H, Acharya R. Quality of care in sterilization services at the public health facilities in India: A multilevel analysis. PLoS One [Internet]. 2020;15(11). Available from: 10.1371/journal.pone.0241499

19. Bansal A, Dwivedi LK. Sterilization regret in India: Is quality of care a matter of concern? Contracept Reprod Med [Internet]. 2020;5(1):13. Available from: 10.1186/s40834-020-00115-8

20. Pulla P. Why are women dying in India’s sterilization camps? BMJ [Internet]. 2014;349. Available from: 10.1136/bmj.g7509

21. Achyut P, Nanda P, Khan N, Verma R. Quality of care in provision of female sterilization in Bihar: A Summary Report. 2014.

22. Kumar A, Gautam A, Dey A, Saith R, Uttamacharya, Achyut P, et al. Infection prevention preparedness and practices for female sterilization services within primary care facilities in Northern India. BMC Health Serv Res. 2019 Dec;20(1):1.

23. Bhatnagar S. Risk of ectopic pregnancy following tubectomy. Indian J Med Res. 1982 Jan;75:47–9.

24. Manesh AO, Sheldon TA, Pickett KE, Carr-Hill R. Accuracy of child morbidity data in demographic and health surveys. Int J Epidemiol [Internet]. 2008 Feb 1;37(1):194–200. Available from: 10.1093/ije/dym202

25. Acharya R, James KS, Singh SK, Saggurti N. Demographic and health surveys and its quality in India. Vol. 24, SSM - population health. England; 2023. p. 101498.

